# Serum, Cell-Free, HPV-Human DNA Junction Detection and HPV Typing for Predicting and Monitoring Cervical Cancer Recurrence

**DOI:** 10.1101/2024.09.16.24313343

**Authors:** Anne Van Arsdale, Olga Mescheryakova, Sonia Gallego, Elaine C. Maggi, Bryan Harmon, Dennis Y.S. Kuo, Koenraad Van Doorslaer, Mark H. Einstein, Brian J. Haas, Cristina Montagna, Jack Lenz

## Abstract

Almost all cervical cancers are caused by human papillomaviruses (HPVs). In most cases, HPV DNA is integrated into the human genome. We found that tumor-specific, HPV-human DNA junctions are detectable in serum cell-free DNA of a fraction of cervical cancer patients at the time of initial treatment and/or at six months following treatment. Retrospective analysis revealed these junctions were more frequently detectable in women in whom the cancer later recurred. We also found that cervical cancers caused by HPV types outside of phylogenetic clade α9 had a higher recurrence frequency than those caused by α9 types in both our study and The Cancer Genome Atlas cervical cancer database, despite the higher prevalence of α9 types including HPV16 in cervical cancer. Thus, HPV-human DNA junction detection in serum cell-free DNA and HPV type determination in tumor tissue may help predict recurrence risk. Screening serum cell-free DNA for junctions may also offer an unambiguous, non-invasive means to monitor absence of recurrence following treatment.

## Introduction

The vast majority of cervical cancers are caused by human papillomaviruses (HPVs), and they remain a significant cause of cancer deaths. Despite the availability of HPV vaccines and effective procedures for detecting and treating pre-invasive cervical lesions, they rank as the fourth most common cancer in women globally^1^. In the United States, an estimated 13,820 new cases of cervical carcinoma are projected for 2024, resulting in 4,360 deaths. The 5-year overall survival (OS) rate across all stages is 66%, with rates dropping to 58% for loco-regional spread and 17% for distant metastasis^2^. Early-stage invasive cervical cancers (FIGO IA1-IB2) are typically treated with surgical excision as a standard of care. However, for patients with advanced stages (FIGO IB3-IVB), surgery is not beneficial, and first line therapy entails cisplatin- radiosensitization combined with ionizing radiation plus brachytherapy, with recurrence risk increasing with stage^3^. An enhanced ability to stratify women during initial care by identifying those at highest risk of treatment failure may improve patient care.

HPV genomes replicate as circular, extra-chromosomal, double-stranded DNAs of about 8 kbp. However, in the vast majority of cervical cancers, HPV DNA is integrated into human DNA, either within a chromosome or as an HPV-human heterocatemeric, extra-chromosomal DNA^4,5,6,7^.

Each junction between HPV and human DNAs is an unambiguous, patient tumor-specific identifier of clonally expanded tumor cells derived from the cell that initially underwent the integration event.

Therefore, we investigated whether HPV-human DNA junctions could be detected circulating in serum cell-free DNA (cfDNA), and whether they could be used to assess residual disease following initial cervical cancer treatment.

## Results

### Detection of HPV-human DNA junctions in cervical cancer patient tumors

HPV-human DNA junctions were retrospectively assessed using archived samples from 16 cervical cancer patients (12 squamous carcinomas, 4 adenocarcinomas) with available frozen tissue and at least one serum sample obtained during initial care. (**Figure 1A, 1B)**. Eight patients had locally advanced cervical cancer (FIGO IB3-IIIC1r), while eight had surgically resectable, early-stage cancers (FIGO IB1-IB2). HPV-human DNA junctions were identified using DNA isolated from tissue biopsies or surgical excisions by hybridization capture for full genomes of 225 HPV types followed by Illumina sequencing (HC+SEQ) and computational identification of junctions using CTAT-Virus Integration Finder^8^ (**Table 1**). This approach also enabled the unambiguous identification of HPV types and genome coverage of each specific type, thus allowing inference of the structures of the integrated viral DNA segments in the tumor genomes (**Figure 1C, 1D, 1E**). This included identification of the lower prevalence types HPV70 and HPV73 in one patient each. In most of the tumors, the inserted HPV DNA comprised a subgenomic fragment of the viral genome (**Figure 1C, 1D**). Complete viral segment sequences determined by HC+SEQ also allows identification of individual, tumor-specific differences compared to HPV type reference sequences at single nucleotide resolution. Junction-PCRs using tumor-specific primers, one near the end of the integrated HPV DNA segment, the other in the flanking human genome sequences (**Methods**) confirmed the HPV-human junctions in the cognate tumor tissue DNAs (data not shown).

**Figure 1.**
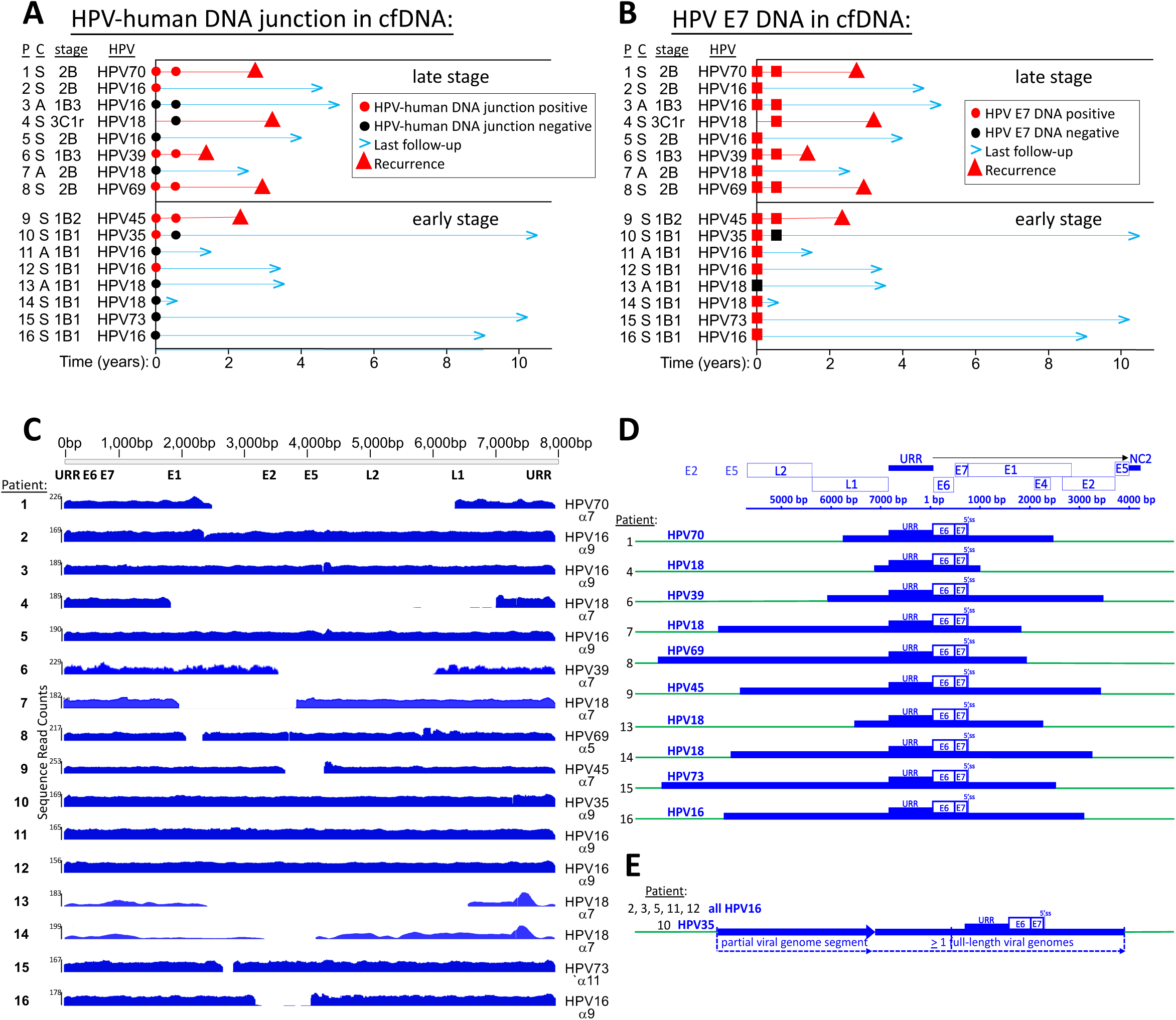
Time course characterizations of the 16 patients in the cohort with individual patients (P), type of cancer (C) either squamous carcinoma (S) or adenocarcinoma (A), diagnostic stage (stage), and HPV type identified in the tumor tissue (HPV). **Panel A** shows analysis of HPV- human DNA junctions in serum cfDNA at initial examination and at the 6-month post-treatment follow-up. **Panel B** shows detection of the cognate HPV E7 DNA in serum cfDNA at the same time points. Descriptions of the symbols are in the box included in each panel. **Panel C** shows the Integrated Genome Viewer (IGV) ^31^ plots of sequence reads obtained by HPV DNA hybridization capture for each sample as a function of the standard HPV genome numbering system^22^. **Panel D** shows the integrated HPV genomic structures for the ten tumors that contained only subgenomic segments of the viral genome. Viral DNAs were centered on the URR-E6-E7 stretch of the viral genomes. HPV open reading frame structure is shown at the top. The 5’ splice site (5’ss) shown in the viral genome is just inside the 5’ end of the E1 open reading frame. As ORF sizes and precise positions vary slightly among HPV types, integrated DNA segments may be slightly off-scale. Some of the HPV DNA insertions had more than two junctions with human DNA clustered within stretches up to tens of kilobase pairs of the human genome, a well-established phenomenon most likely due to instability of inserted HPV DNA including extra-chromosomal circularization of HPV-human DNA heterocatemer segments, and nearby reintegration of such structures^4–6,24–28^. In those instances, the junctions shown were evident as read gap borders in Panel C, and they also had the largest number of PCR-confirmed, hybridization capture + sequencing reads. **Panel E** shows the presumed genomic structures of integrated HPV DNAs in six tumors containing viral segments that were longer than unit length. All were from phylogenetic clade α9, five HPV16 and one HPV35. Arrows show the viral 5’ to 3’ transcriptional orientation of the HPV protein coding strands.

**Table 1.**
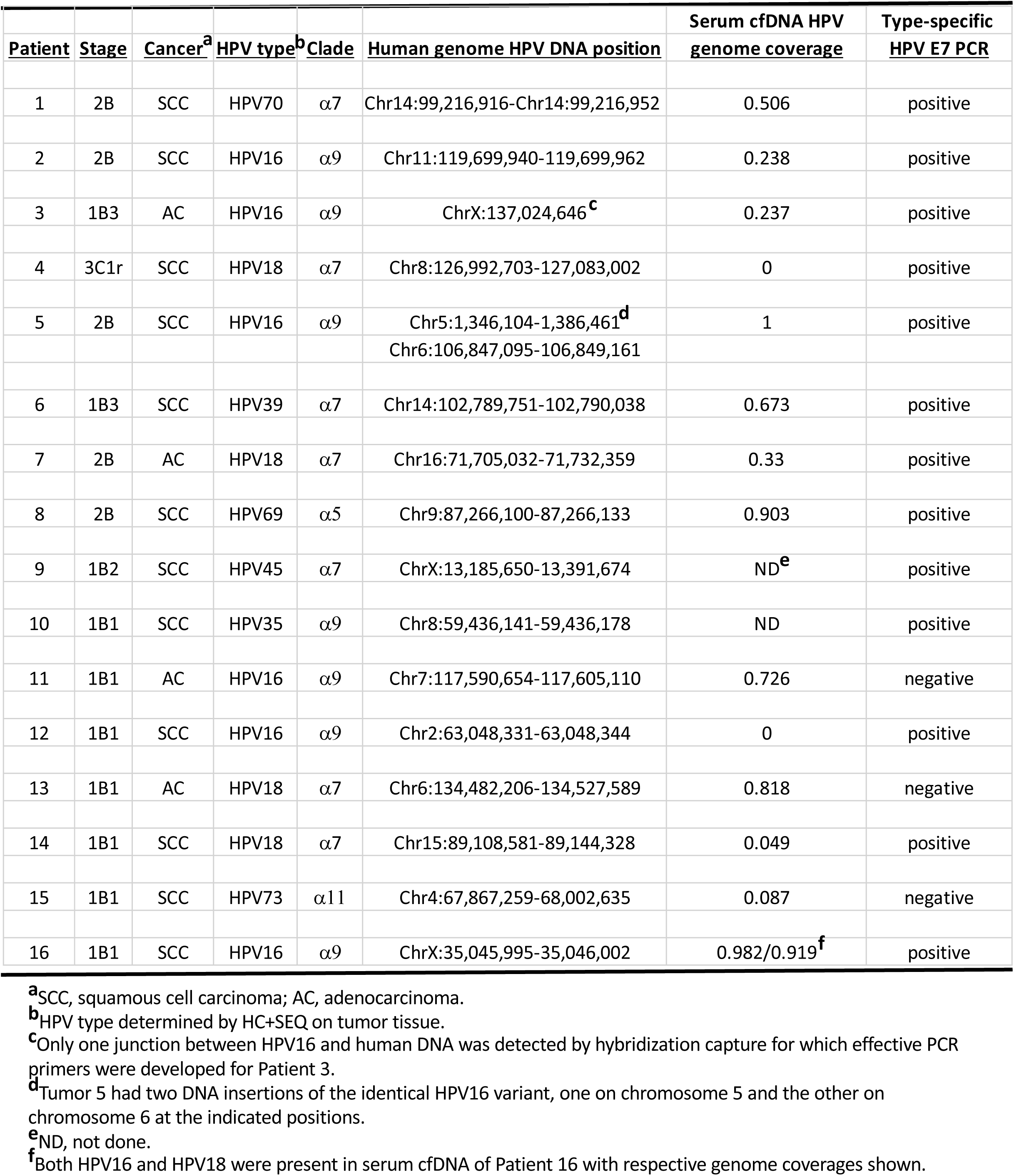
HPV types, viral phylogenetic clades, human genome integration sites, serum cfDNA coverage by HC+SEQ, and serum cfDNA type-specific E7 PCR.

| <u>Patient</u> | <u>Stage</u> | <u>Cancer<sup>a</sup></u> | <u>HPV type<sup>b</sup></u> | <u>Clade</u> | <u>Human genome HPV DNA position</u> | <u>Serum cfDNA HPV genome coverage</u> | <u>Type-specific HPV E7 PCR</u> |
| --- | --- | --- | --- | --- | --- | --- | --- |
| 1 | 2B | SCC | HPV70 | $\alpha 7$ | Chr14:99,216,916-Chr14:99,216,952 | 0.506 | positive |
| 2 | 2B | SCC | HPV16 | $\alpha 9$ | Chr11:119,699,940-119,699,962 | 0.238 | positive |
| 3 | 1B3 | AC | HPV16 | $\alpha 9$ | ChrX:137,024,646<sup>c</sup> | 0.237 | positive |
| 4 | 3C1r | SCC | HPV18 | $\alpha 7$ | Chr8:126,992,703-127,083,002 | 0 | positive |
| 5 | 2B | SCC | HPV16 | $\alpha 9$ | Chr5:1,346,104-1,386,461<sup>d</sup><br>Chr6:106,847,095-106,849,161 | 1 | positive |
| 6 | 1B3 | SCC | HPV39 | $\alpha 7$ | Chr14:102,789,751-102,790,038 | 0.673 | positive |
| 7 | 2B | AC | HPV18 | $\alpha 7$ | Chr16:71,705,032-71,732,359 | 0.33 | positive |
| 8 | 2B | SCC | HPV69 | $\alpha 5$ | Chr9:87,266,100-87,266,133 | 0.903 | positive |
| 9 | 1B2 | SCC | HPV45 | $\alpha 7$ | ChrX:13,185,650-13,391,674 | ND<sup>e</sup> | positive |
| 10 | 1B1 | SCC | HPV35 | $\alpha 9$ | Chr8:59,436,141-59,436,178 | ND | positive |
| 11 | 1B1 | AC | HPV16 | $\alpha 9$ | Chr7:117,590,654-117,605,110 | 0.726 | negative |
| 12 | 1B1 | SCC | HPV16 | $\alpha 9$ | Chr2:63,048,331-63,048,344 | 0 | positive |
| 13 | 1B1 | AC | HPV18 | $\alpha 7$ | Chr6:134,482,206-134,527,589 | 0.818 | negative |
| 14 | 1B1 | SCC | HPV18 | $\alpha 7$ | Chr15:89,108,581-89,144,328 | 0.049 | positive |
| 15 | 1B1 | SCC | HPV73 | $\alpha 11$ | Chr4:67,867,259-68,002,635 | 0.087 | negative |
| 16 | 1B1 | SCC | HPV16 | $\alpha 9$ | ChrX:35,045,995-35,046,002 | 0.982/0.919<sup>f</sup> | positive |

<sup>a</sup>SCC, squamous cell carcinoma; AC, adenocarcinoma.
<sup>b</sup>HPV type determined by HC+SEQ on tumor tissue.
<sup>c</sup>Only one junction between HPV16 and human DNA was detected by hybridization capture for which effective PCR primers were developed for Patient 3.
<sup>d</sup>Tumor 5 had two DNA insertions of the identical HPV16 variant, one on chromosome 5 and the other on chromosome 6 at the indicated positions.
<sup>e</sup>ND, not done.
<sup>f</sup>Both HPV16 and HPV18 were present in serum cfDNA of Patient 16 with respective genome coverages shown.

Five of the sixteen patients had tumor recurrences, all occurring at distant metastatic sites outside standard-of-care radiation fields or excised organs. Metastasis biopsy samples were available from all five recurrences, and junction-PCR analysis on DNAs from them confirmed identical junctions to those found in each cognate primary tumor (**Figure 2)**, thus confirming clonal derivation from the primary tumors.

**Figure 2.**
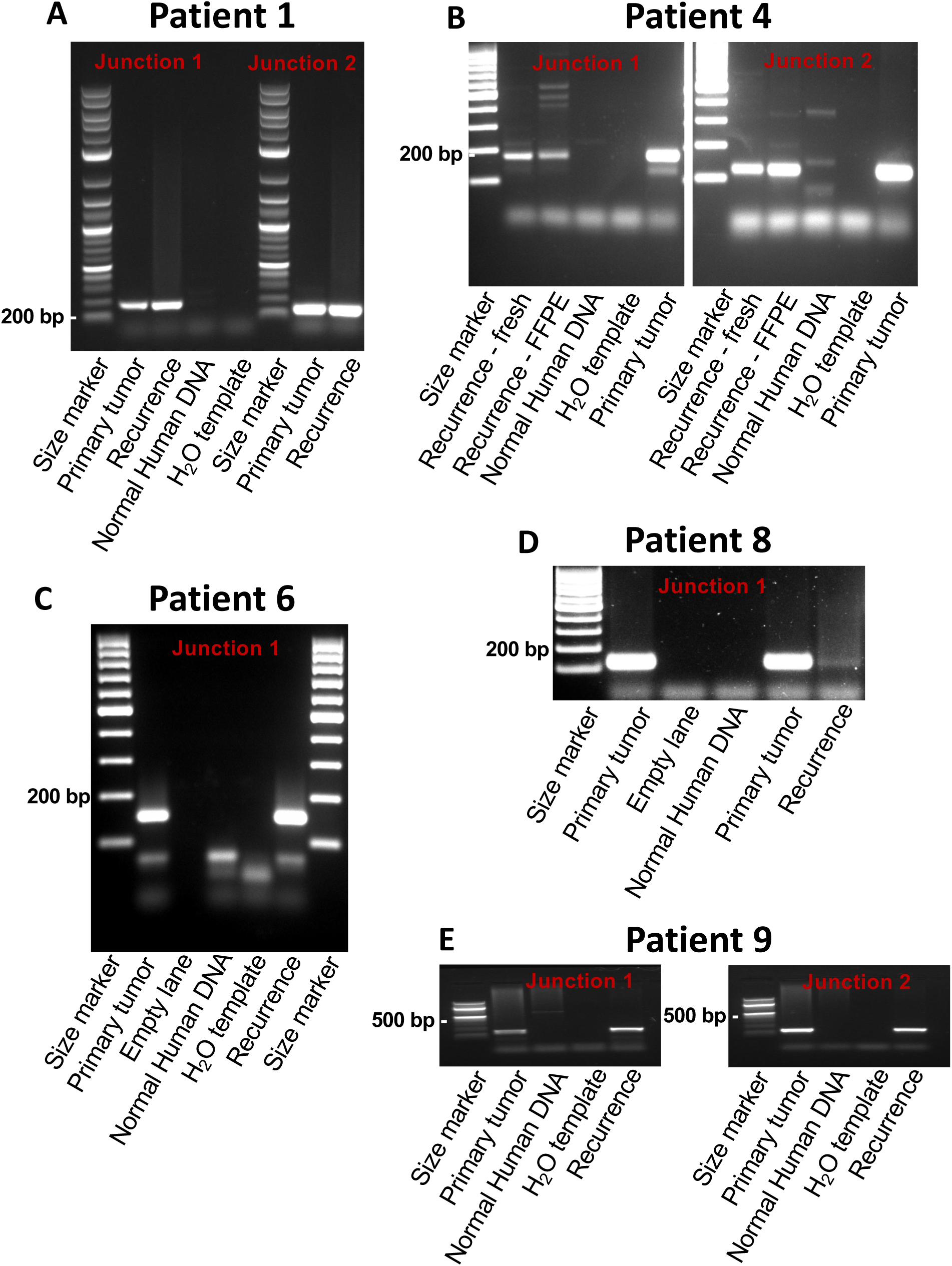
PCR detection of HPV-human DNA junctions in human genomic DNAs isolated from biopsy tissue of the cancer recurrences that occurred in the five indicated patients. PCRs for each patient included a sample of DNA from the recurrence and a positive control using the primary tumor tissue DNA from the individual patients. The junctions in the five recurrences were also detected by hybridization capture plus Illumina sequencing on the recurrence biopsies, which likewise confirmed clonal derivation from the cognate primary tumors.

### Tumor-derived, HPV-human DNA junctions in serum cell-free DNA

The usefulness of junction detection in serum cfDNAs was evaluated using samples obtained during initial care from fifteen of the patients and at six months post-treatment from seven of the patients (**Figure 1A**). Since levels of cfDNA in serum from cancer patients are generally elevated over levels in control individuals^9^, serum DNA concentrations were measured in the samples obtained at initial care. They were significantly higher in women diagnosed with locally advanced cancers stage compared to those with early-stage cancers (Wilcoxon rank sum test, p<0.05) (**Figure 3A**), but no significant difference was observed between those with or without a recurrence (**Figure 3B**). HPV-human DNA junctions in cfDNAs were assessed by HC+SEQ directly on cfDNA and by junction PCR methods, which yielded concordant results except for Patient 2 (**Methods**). Junctions were detected in cfDNA from seven patients, including all cases of recurrences except no baseline sample was available for Patient 4 (**Figure 1A**).

**Figure 3.**
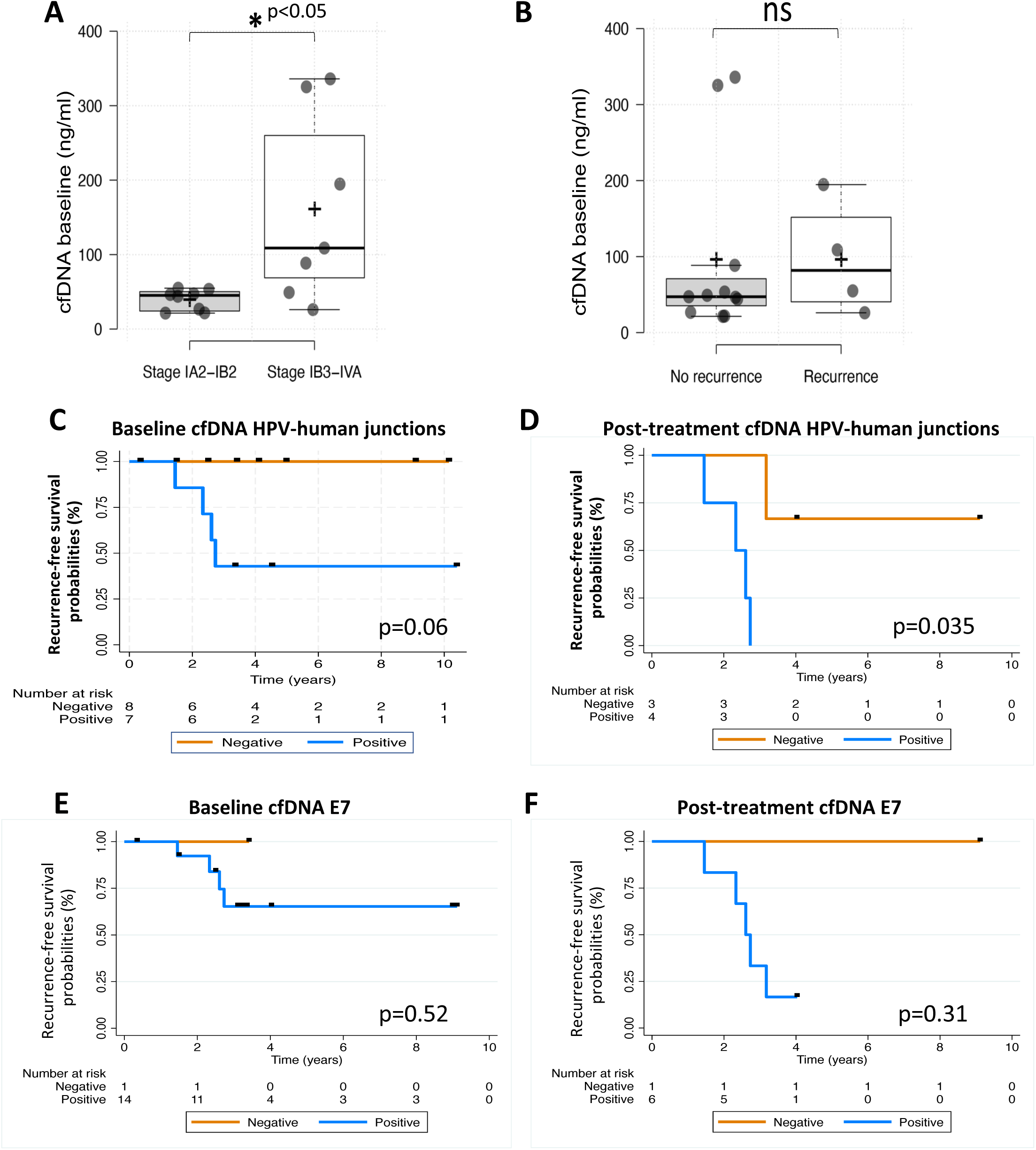
DNA concentration determinations and Kaplan-Meier analyses performed using cfDNA from patients. **Panel A** shows total cfDNA concentrations in early- vs. late-stage cancers. **Panel B** shows the same DNA concentration determinations plotted from patients who did not have a recurrence vs. those that did. **Panels C and D** show Kaplan-Meier analyses of recurrence-free survival in patients where HPV-human DNA junctions were detected in serum vs. those where it was not detected at initial examination (**Panel C**) or at 6-months post- treatment (**Panel D**). **Panels E and F** show Kaplan-Meier analyses of recurrence-free survival in patients where HPV E7 DNA was detected in serum vs. those where it was not detected at initial examination (**Panel E**) or at 6-months post-treatment (**Panel F**). In all four plots, squares show left-censoring at the last patient visits.

### HPV-human DNA junction detection in cfDNA for prediction of recurrence-free survival

In light of HPV-human DNA junctions being unambiguous markers for tumor cell clones containing integrated HPV DNA, the significance of junction detection for predicting recurrence at baseline and six months was assessed by Kaplan-Meier analyses of recurrence-free survival **(Figures 3C, 3D**). Stratified log-rank tests adjusted for stage at diagnosis and left-censored to the dates of last-follow-up showed that detection at 6 months was statistically significant and prognostic for recurrence (p=0.035). Detection at initial treatment was of borderline statistical significance (p=0.06). Due to the small sample size, accurate determination of effect size of detection at either time point with appropriate 95% confidence intervals could not be accomplished using either univariable or multivariable Cox regression modeling. Nonetheless, junction detection in serum cfDNA at 0 and 6 months showed promise as a predictor of tumor recurrence.

HPV DNA detection alone in cfDNA of cervical cancer patients has demonstrated value for helping predict tumor recurrence risk^10^. The E7 gene is crucial for HPV tumorigenesis^11^, and the URR-E6-E7 segment was present in all the tumors (**Figure 1D and 1E**). However, while circulating tumor DNA (ctDNA) is the predominant component of cfDNA in cancer patients, HPV DNA from non-tumor tissues could also be present in cfDNA^12,13^. Therefore, HPV type-specific E7 DNA detection in cfDNA was also evaluated (**Figures 1B**). Among available samples, 14/15 (93.3%) baseline serum cfDNAs and 6/7 post-treatment sera, including all five from patients with recurrences, tested positive for HPV type-specific E7 DNA by HC+SEQ and/or E7 gene PCR (**Table 1**). No HPV DNA was detected by PCR or HC+SEQ in serum cfDNA from Patient 13, where HPV18 DNA was integrated in the cognate tumor. Kaplan-Meier analysis at baseline and six months post-treatment using stratified log-rank tests for HPV E7 positivity, adjusted for stage, did not significantly associate with recurrence at either time point (p=0.52 and p=0.31, respectively) (**Figure 3E and 3F**). Due to the small sample size, accurate effect size determination with 95% confidence intervals using univariable or multivariable Cox regression modeling was not feasible.

Interestingly, Patient 16 serum cfDNA tested positive for both HPV16 and HPV18 DNA by HC+SEQ with genome coverage fractions of 0.982 and 0.919, respectively, and by E7 PCRs (**Figure 4**, **Table 1**). In contrast, the primary tumor tested positive only for HPV16 DNA by HC+SEQ and E7 PCR (**Table 1 and Figure 4A, 4B**). Moreover, the HPV16 DNA detected in serum cfDNA had different mutations relative to a reference HPV16 genome than the HPV16 DNA in the tumor (**Figure 4C**). These results implied that the cervical cancer in this patient was caused by the HPV16 variant detected in the tumor tissue, and that neither the HPV16 nor the HPV18 that were detected in the serum cfDNA were present in the tumor at the time of initial treatment and presumably derived from an alternative source. Moreover, no junctions with human DNA were detected with either of the HPV DNAs in serum cfDNA, and HPV16 sequence reads in cfDNA were continuous across the viral genome at the positions of the junctions with human DNA in the cancer. These observations suggested that the patient was coinfected with both the HPV16 and HPV18 found in serum cfDNA at the time that the baseline sample was obtained, and that there was no evidence for a causative, tumorigenesis role by either those HPVs. In summary, these results indicate that HPV DNA in serum cfDNA may arise from unclear tissue sources, and that HPV-human DNA junctions provide a more rigorous, tumor-specific means for detecting the presence of tumor cells than detection of just HPV DNA, at least in some patients.

**Figure 4.**
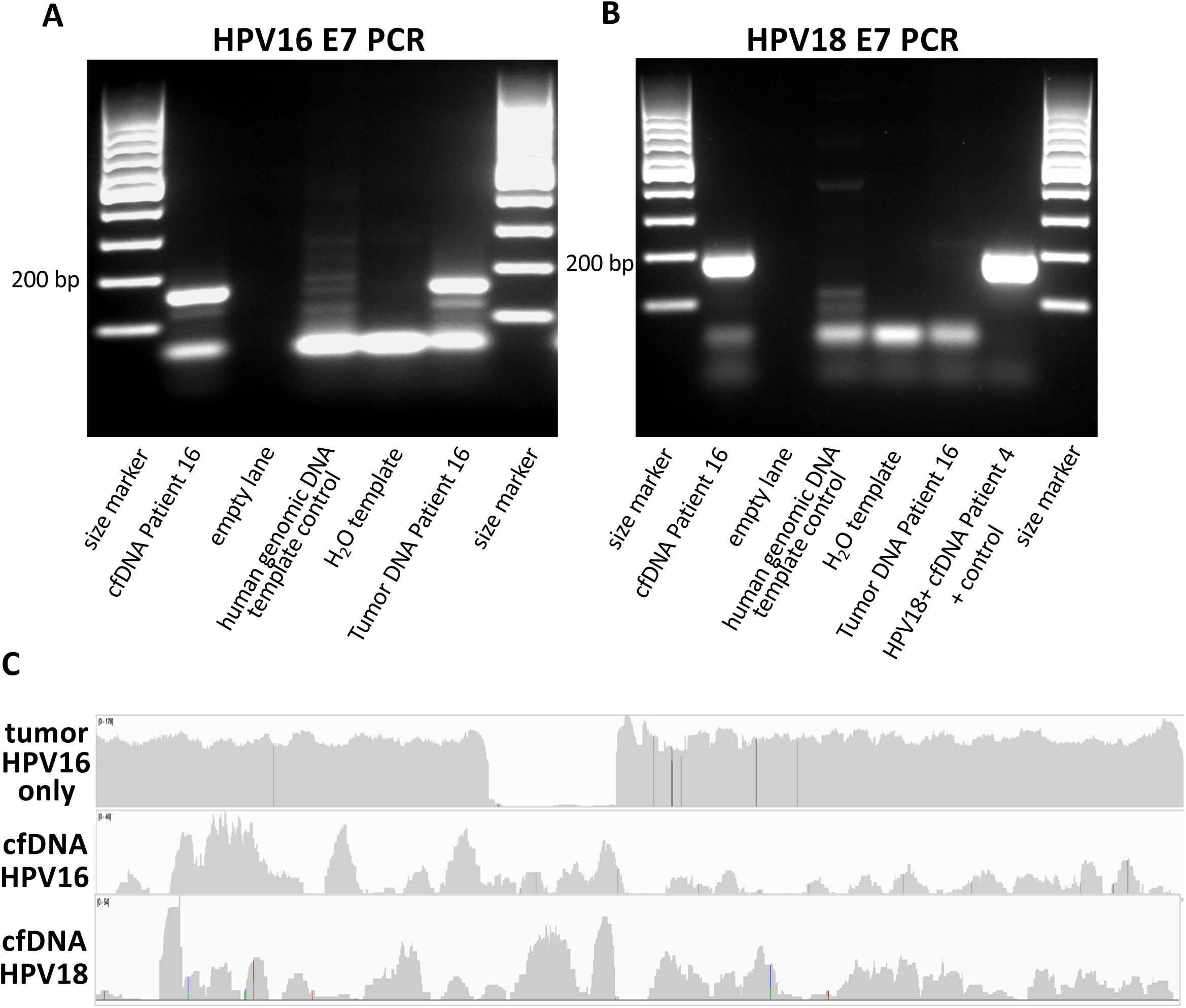
Analyses of HPV DNAs in Patient 16. **Panel A** shows PCR analysis for HPV16 E7 DNA in serum cfDNA plus DNA from the primary tumor as a positive control. **Panel B** shows PCR analysis for HPV18 E7 DNA in serum cfDNA plus DNA from the primary tumor as a positive control. **Panel C** shows the IGV plots of sequence reads following HPV DNA hybridization capture from the primary tumor and cfDNA. Sequence reads for HPV16 and HPV18 in serum cfDNA were plotted separately.

### HPV phylogenetic clade type as a risk factor for cancer recurrence

Unexpectedly, all five recurrences in this cohort involved HPV types from phylogenetic clade α7 (HPV18, HPV39, HPV45, HPV70) or α5 (HPV69), with none involving clade α9 types (HPV16, HPV35) (**Figure 1A**). Patients with cervical cancers containing clade α7 HPV types have inferior survival versus those with clade α9^14,15^. Therefore, HPV clade types were also assessed as an independent prognostic factor for recurrence for the larger cervical cancer dataset in The Cancer Genome Atlas (TCGA) using Kaplan-Meier analysis of clinical information and tumor tissue sequences (**Tables 2 and 3**, **Figure 5**). HPV types were determined using CTAT-Virus Integration Finder^8^ analysis of TCGA sequencing data, revealing 214 tumors (69.5%) with clade α9 types, 83 (26.9%) with other clade types (α5, α6, α7, or α11) , and 11 (3.6%) being HPV-negative. Clinicopathologic variables known to be *a priori* prognostic factors were extracted from cBioPortal^16^ ^17^, including age, histology, and stage at diagnosis, with N=297 patients having complete data. Comparisons of HPV clade α9 with non-clade-α9 (α5/α6/α7/α11) showed no significant baseline differences in these variables (**Table 2**). However, patients with non-α9 clades exhibited inferior recurrence-free survival compared to those with HPV clade-α9 tumors based on a log-rank test stratified for histology and stage (p=0.03). A multivariable Cox regression model including age (years), histology (squamous/adenocarcinoma) and stage (I, II, III, IV) (**Table 3**) was performed to account for these *a priori* variables, and women with HPV clades other than α9 showed inferior recurrence-free survival in comparison to HPV clade-α9 tumors (Hazard Ratio (HR)=2.48, 95% CI: 1.34-4.49, p=0.004) after adjusting for all variables (**Figure 5**). The effect of HPV clade on recurrence-free survival was second only to stage IV disease (HR: 4.40, 95% CI: 1.89-10.18, p=0.001) in this adjusted analysis (**Table 3**). In summary, these results show that non-α9 clade HPV types are a risk factor for tumor recurrence following current standards of care.

**Figure 5.**
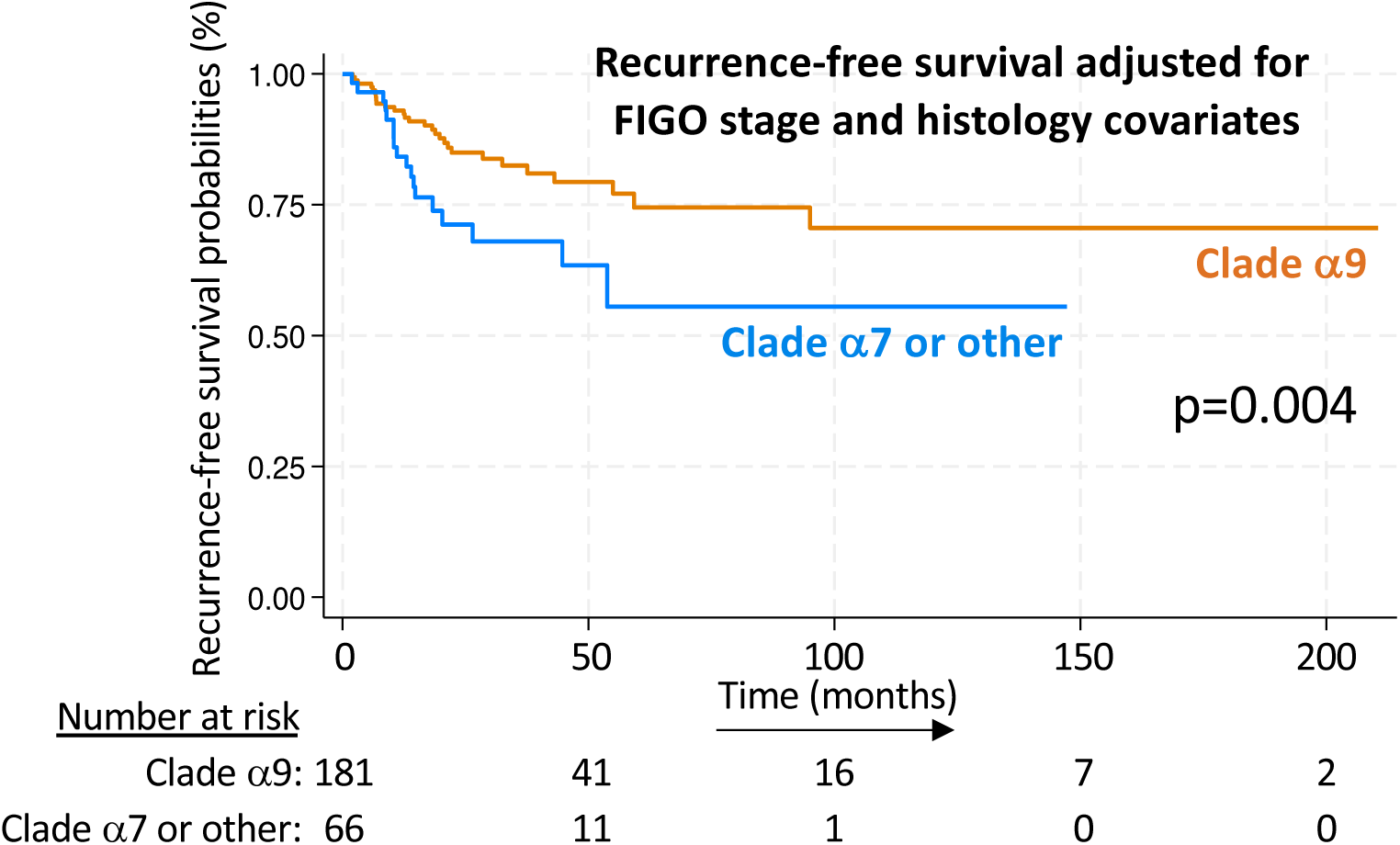
Kaplan-Meier recurrence-free survival plot of 297 TCGA cervical cancer patients.

**Table 2.** *A priori* clinicopathologic variables of the TCGA cohort by HPV phylogenetic clade.

| Variable | Cohort (n=297) | Clade 9 (n=214) | Clade 7 or other (n=83) | p-value |
| --- | --- | --- | --- | --- |
| Age (years) | 47.8 (13.6) | 48.6 (13.8) | 45.8 (13.0) | 0.12 |
| **Histology** |  |  |  |  |
| Squamous Cell | 254 (84.4) | 183 (85.9) | 67 (83.8) | 0.71 |
| Adenocarcinoma | 47 (15.6) | 30 (14.1) | 13 (16.2) |  |
| **Stage at Dx** |  |  |  | 0.57 |
| Stage I | 156 (54.6) | 113 (54.6) | 43 (54.4) |  |
| Stage II | 65 (22.7) | 44 (21.3) | 21 (26.6) |  |
| Stage III | 46 (16.1) | 34 (16.4) | 12 (15.2) |  |
| Stage IV | 19 (6.6) | 16 (7.7) | 3 (3.8) |  |

**Table 3.** Multivariable Cox proportional hazard regression model for recurrence-free survival.

| Variable | Multivariable Hazard Ratio | 95% CI | p-value |
| --- | --- | --- | --- |
| **Clade** |  |  |  |
| Clade 9 | 1.00 | -- | -- |
| Clade 7 | 2.48 | 1.34-4.49 | 0.004 |
| **Age (years)** | 1.01 | 0.99-1.04 | 0.28 |
| **Histology** |  |  |  |
| Squamous | 1.00 | -- | -- |
| Adenocarcinoma | 1.08 | 0.51-2.28 | 0.84 |
| **Stage at Dx** |  |  |  |
| Stage I | 1.00 | -- | -- |
| Stage II | 0.63 | 0.28-1.41 | 0.26 |
| Stage III | 0.43 | 0.13-1.46 | 0.18 |
| Stage IV | 4.40 | 1.89-10.18 | 0.001 |

## Discussion

The results presented here show that HPV-human junction detection in serum cfDNA and that HPV type determination in tumor tissue at the time of initial cervical cancer treatment might be useful to help predict the likelihood of cervical cancer recurrence. In particular, they also show that junction detection at six-months post-treatment may be useful for unambiguous detection of residual tumor tissue. Detection of HPV DNA alone in cfDNA is useful for predicting relapse^10 18^. However, the detection of HPV’s different than the one integrated in the cfDNA of Patient 16 here (**Figure 4**) illustrates the potential for occasional, misleading results, and HPV DNA has been reported in blood of healthy donors and women with documented preinvasive cervical dysplasia up to 50% of the time^12,13^. These are consistent with the possibility that HPV DNA in cfDNA may occasionally arise from non-tumor tissue sources, thus emphasizing the potential advantage of HPV-human DNA junctions for assessing residual disease. Our results also suggest that HPV-human DNA junctions could be tested in other types of HPV-induced cancers.

HPV-human DNA junctions in cfDNA offer the advantage of unequivocal origin from the cancer cell clones in patients. Long-term prospective studies are required to confirm the findings here, and the potential for improved recurrence risk prediction by HPV-human DNA junction detection in serum emphasizes the need to develop improved upfront therapies for women at risk for cervical cancer relapse. In addition, junction detection in serum cfDNA potentially offers a non-invasive means for rigorous, longitudinal monitoring of disease recurrence in patients following cervical cancer treatment.

Identification of non-α9 clade HPV types (at least α5, α6, α7, and α11) as a risk factor for recurrence in the TCGA/cBioPortal data^16^ ^17^ and in the small retrospective study presented here establish that HPV clade type specifically affects cervical cancer recurrence *per se* following treatment as well as survival, and may reflect the same underlying molecular genetic properties of these HPV types. These effects contrast with the high prevalence of α9 HPV16 (∼50%) in cervical cancer and the correlation between HPV16 and progression of preinvasive, cervical intraepithelial neoplasia^19^ ^20^. They also highlight a potential role for precise HPV typing in cervical cancer management using multiplexed HPV-typing by hybridization capture plus sequencing methods like HC+SEQ as used here to facilitate clade assignment. These approaches are technically comparable to clinically-utilized assays like exon capture, and inclusion of all known HPV types also enables broad diagnostic specificity plus tumorigenicity assessment of rare HPV types. Again, the potential for improved recurrence risk prediction emphasizes the need to develop improved upfront therapies for women at risk for cervical cancer relapse.

In summary, HPV-human DNA junction detection in cfDNA and HPV type determination at the time of initial treatment offer promise for predicting and monitoring cervical cancer recurrence.

## Methods

### Patients, sample collection and nucleic acid preparation

All procedures were approved by the Internal Review Board of the Albert Einstein College of Medicine at Einstein-Montefiore. Tumors from women diagnosed with invasive cervical carcinoma were collected as part of an ongoing tissue collection protocol of the Department of Obstetrics & Gynecology and Women’s Health at the Albert Einstein College of Medicine (IRB# 2009-265). 16 cervical cancer patients were chosen retrospectively based on availability of frozen tumor tissue plus an archived serum sample from time of initial treatment and/or 6 months post-treatment. All patients receiving adjuvant or curative radiotherapy completed their course within 8 weeks. Diagnosis was by standard-of-care histopathology on biopsies and included 12 (75%) squamous cell carcinomas and 4 (25%) adenocarcinomas (**Figure 1**, **Table 1**), which is reflective of published epidemiological prevalence data.

From all samples, 10 μM sections of the frozen tissue were cut for H&E confirmation of the presence of viable tumor tissue and presence of the diagnosed histologic subtype as reviewed by a board-certified gynecologic pathologist (Bry.H.). DNA was extracted from sections of the frozen tissue using the QIAamp DNA Mini Kit (Qiagen) stored at −80°C until use. Serum cfDNA was extracted from samples banked at date of diagnosis (baseline) and/or at 6 months post-treatment completion. Serum was also collected the five patients that suffered a recurrence at the time of the recurrence. Qiagen QIAamp Circulating Nucleic Acid kits were used to extract serum cfDNA. Extracted DNAs were subjected to two separate serial dilutions, and DNA concentrations were quantified using the Qubit Fluorometric Quantification method (Thermo Fisher Scientific) and DNA integrity was assessed using the Bioanalyzer capillary electrophoresis system (Agilent).

### HPV DNA hybridization capture with short read Illumina sequencing (HC+SEQ) and data analysis

HPV DNA including fragments containing HPV-human DNA junctions were enriched using a DNA hybridization-capture panel containing the genomes of 225 HPV types, followed by Illumina sequencing. Sequence reads were aligned to a custom-assembled sequence file comprising the human and 225 HPV genomes to identify HPV types, assess viral genome coverage, and identify HPV-human DNA junctions.

Targeted enrichment of HPV DNA was performed using custom hybridization capture probes homologous to the full-length genomes of 225 different HPV types as previously described^5,21^. Briefly, 1 µg of tumor genomic DNA was mechanically fragmented to 200 bp for short-range sequencing (Covaris, Woburn, MA), and Illumina adaptors were ligated at each end using the KAPA Hyper Prep kit per manufacturer instructions. Libraries were then hybridized to the biotinylated HPV oligonucleotide probes specific for each of the 225 HPV types for 72 h using the Roche target enrichment protocol (Roche Nimblegen SeqCap EZ System, Basel, Switzerland) following manufacturer instructions. After library purification, the DNA was sequenced on one Illumina Novaseq lane (Illumina, San Diego, CA) using the paired end, 150 bp sequencing mode.

The sequencing reads were next adaptor-cleaned, and those that passed QC were de-duplicated, and paired end reads were aligned to a custom human (GRCh38/hg38) plus HPV reference genome containing 225 HPV types from the Papillomavirus Episteme^22^ using the STAR Aligner^23^. Average sequencing coverage of the sample typed HPV genome after de-duplication was calculated based on the mapped, on-target, 150 bp reads. HPV-human junction fragments were computationally identified using CTAT-Virus Integration Finder that specifies a chimeric read analysis to define integration sites^8^. When integrated HPV DNAs were of subgenomic length, the edges of the HPV genome coverage plots (**Figure 1C**) uniformly corresponded to junctions with human DNA (**Figure 1D**). In some tumors, more than two junctions were identified during HC+SEQ that each clustered within a single segment of the human genome, a previously described phenomenon^24–28^ suggesting that integrated HPV DNA segments undergo rearrangements to some extent at least in some lesions. The junctions illustrated and used for PCR analyses here represent those with the highest numbers of HC+SEQ read counts. The other junctions presumably represent subclonal variants and may be similarly useful for the types of junction analyses described in this study. HC+SEQ using serum cfDNA was performed similarly except that the amounts of starting material were lower (5 to 100 ng). No additional amplification was performed beyond the KAPA Hyper Prep step and the final capture libraries.

### HC+SEQ sequence data availability

HC+SEQ sequence reads are available (SUB14649435) at the Sequence Read Archive (SRA) of the National Library of Medicine, National Institute of Health.

### HPV type and human DNA junction analysis by PCR and Sanger sequencing

PCR was also used to confirm the human genome junctions and HPV types detected by HC+SEQ. PCR primers to detect the HPV-human DNA segments unique to each individual tumor/cfDNA sample (junction PCRs) were designed to flank each side of the junctions obtained from HC+SEQ split reads with one primer in the HPV sequences and the other in the flanking human sequences. We were unable to design primers that successfully amplified the junctions for patient 2, even though they could be detected in the cfDNA by HC+SEQ. HPV type was also assessed using PCR primers specific for the E7 genes of each HPV type, as it was retained in all tumor samples from the cohort (**Figure 1**). Primers and amplicon size for junctions positive by PCR are in **(Supplementary Table 1)**. PCR reactions were performed using GoTaq Green (Promega) using a standard of 35 cycles of 94°C for 30 sec, 60°C for 30 sec, and 72°C for 45 sec and adjusted if needed based on individual primer Tm. PCR products were resolved using a 2% agarose gel and extracted using the QIAquick® Gel Extraction Kit (Qiagen) for Sanger sequencing.

### Statistical Analysis and TCGA dataset

Detailed medical records were abstracted to obtain data regarding age at diagnosis, histological subtype, stage, and treatment related variables (**Supplementary Table 2**). Histologic classification was designated either squamous cell carcinoma or adenocarcinoma of the cervix. Stage was determined by the 2018 International Federation of Gynecology and Obstetrics (FIGO) staging criteria^29^. Treatment related variables included: 1) primary surgical intervention which consisted of radical hysterectomy, bilateral salpingo-oophorectomy, and pelvic lymph node dissection, 2) radiation therapy delivered as external beam and/or brachytherapy as either for primary curative intent or as adjuvant therapy based on Sedlis criteria^30^ and 3) systemic cisplatin chemotherapy administered as a radiosensitizer in either primary curative intent or adjuvant setting. Based on biopsy, Patient 9 was diagnosed with stage 1B2, but based on histopathological analysis of the surgically excised tissue, additional high-risk features were noted including positive parametria and positive pelvis lymph nodes, and thus also received chemo-radiation therapy. HPV type was determined based on HC+SEQ HPV typing and PCR confirmation. Patients were stratified into HPV phylogenetic clades for the viral types in this cohort as follows: HPV clade α7 (types 18, 39, 45 and 70), HPV clade α9 (HPV types 16 and 35), and HPV clades “other” (HPV types 69 and 73, clades α5 and α11, respectively).

The primary exposures of interest were 1) HPV-human DNA junction positivity by either HC+SEQ or PCR and 2) HPV E7 positivity by either HC+SEQ or PCR. The primary outcome of interest was time to recurrence. Recurrence-free survival (RFS) was calculated from the date of diagnosis to the date of first recurrence. Patients were left-censored at the date of last follow up. This included patients that were lost to follow up, which was assumed non-informative, and patients that did not experience a recurrence event.

Demographic and disease characteristics by recurrence status were described using means and standard deviation for normally distributed continuous variables, medians and interquartile ranges (IQR) for non-normally distributed variables, and frequencies and percentages for categorical variables. Differences in these variables were assessed using the Student’s t-test, Wilcoxon test, and Fisher’s Exact test, respectively. Recurrence-free survival was summarized by HPV-human DNA junction or HPV E7 baseline positivity, and by HPV-human junction or HPV E7 at 6 months post-treatment using Kaplan-Meier survival curves. Differences were assessed using the stratified log-rank test, stratifying on the *a priori* variables histology and stage, which are both independently associated with disease recurrence.

To evaluate whether HPV type was associated with recurrence in the cervical cancer dataset of The Cancer Genome Atlas, we downloaded the RNA-sequencing, whole genome sequencing (WGS), and/or whole exome sequencing (WES) data for 302 cervical cancer samples from TCGA^16^. HPV typing was determined using CTAT-Virus Integration Finder developed by our group^8^. Sample HPV type was determined based on support with at least 50 mapped reads, >0.10 fraction genome coverage. Based on the inferred HPV type, samples were categorized into HPV clades α9, and “α7 plus other” as described above. RFS and demographic variables including age, histology, and stage at diagnosis were extracted from CBioportal^17^. Survival probabilities for RFS were estimated using the Kaplan-Meier method, and the log-rank test was used to compare RFS by HPV clade. Univariable and multivariable Cox proportional hazard regression analyses were also performed to assess further whether HPV clade was an independent risk factor for RFS. Previously reported risk factors related to RFS included in all models were age, stage, and histology. All analyses were performed using Stata version 18.0 (StataCorp, College Station, TX).

## Data Availability

All sequencing data produced are available online at SRA

https://www.ncbi.nlm.nih.gov/sra

## Acknowledgements

This work was supported by NIH grants CA240580 (CM and JL), CA180922 (Bri.H.), and Montefiore Einstein Cancer Center grant 3A3216. AVA was supported by NIH K08 CA273527 and an American Association of Obstetricians and Gynecologists (AAOFG) Scholar Award.

---

**Supplementary Table 2.** Patient clinical parameters.

| Variable | Cohort (n=16) | No Recurrence (n=11) | Recurrence (n=5) | p-value<sup>a</sup> |
| --- | --- | --- | --- | --- |
| **Age (years)** | 57.1 (13.7) | 52.6 (12.4) | 67 (12.0) | 0.048 |
| **Histology** |  |  |  |  |
| Squamous Cell | 12 (75.0) | 7 (63.6) | 5 (100.0) | 0.25 |
| Adenocarcinoma | 4 (25.0) | 4 (36.4) | 0 (0.0) |  |
| **Stage** |  |  |  | 0.28 |
| Early (IA-IB2) | 8 (50.0) | 7 (63.6) | 1 (20.0) |  |
| LACC (IB3-IVA) | 8 (50.0) | 4 (36.4) | 4 (80.0) |  |
| **HPV by Clade** |  |  |  | 0.013 |
| Clade 7 | 6 (37.5) | 2 (18.2) | 4 (80.0) |  |
| Clade 9 | 8 (50.0) | 8 (72.7) | 0 (0.0) |  |
| Other | 2 (12.5) | 1 (9.1) | 1 (20.0) |  |
| **Baseline cfDNA concentration (ng/mL)** | 49.1 [21.4,336] | 47.2 [21.4, 336] | 81.9 [26.1, 194.7] | 0.49 |
| **Surgery** |  |  |  | 0.30 |
| No | 6 (37.5) | 3 (27.3) | 3 (60.0) |  |
| Yes | 10 (62.5) | 8 (72.7) | 2 (40.0) |  |
| **EBRT + brachy<sup>b</sup>** |  |  |  | 0.25 |
| No | 4 (25.0) | 4 (36.4) | 0 (0.0) |  |
| Yes | 12 (75.0) | 7 (63.6) | 5 (100.0) |  |
| **Total Dose (Gy)** | 69 [60,84] | 60 [60,78] | 81 [60,84] | 0.07 |
| **Chemotherapy** |  |  |  | 0.31 |
| No | 7 (43.8) | 6 (54.6) | 1 (20.0) |  |
| Yes | 9 (56.2) | 5 (45.4) | 4 (80.0) |  |

<sup>a</sup>Students t-test, Wilcoxon test, or Fisher's Exact test were utilized for normally distributed variables, non-normal variables and categorical variables respectively. p<0.05 was considered statistically significant.
<sup>b</sup>External beam radiotherapy (EBRT) and brachytherapy (brachy)

