## Supplementary Tables for "Serum, Cell-Free, HPV-Human DNA Junction Detection and HPV Typing for Predicting and Monitoring Cervical Cancer Recurrence"

### Slide 1
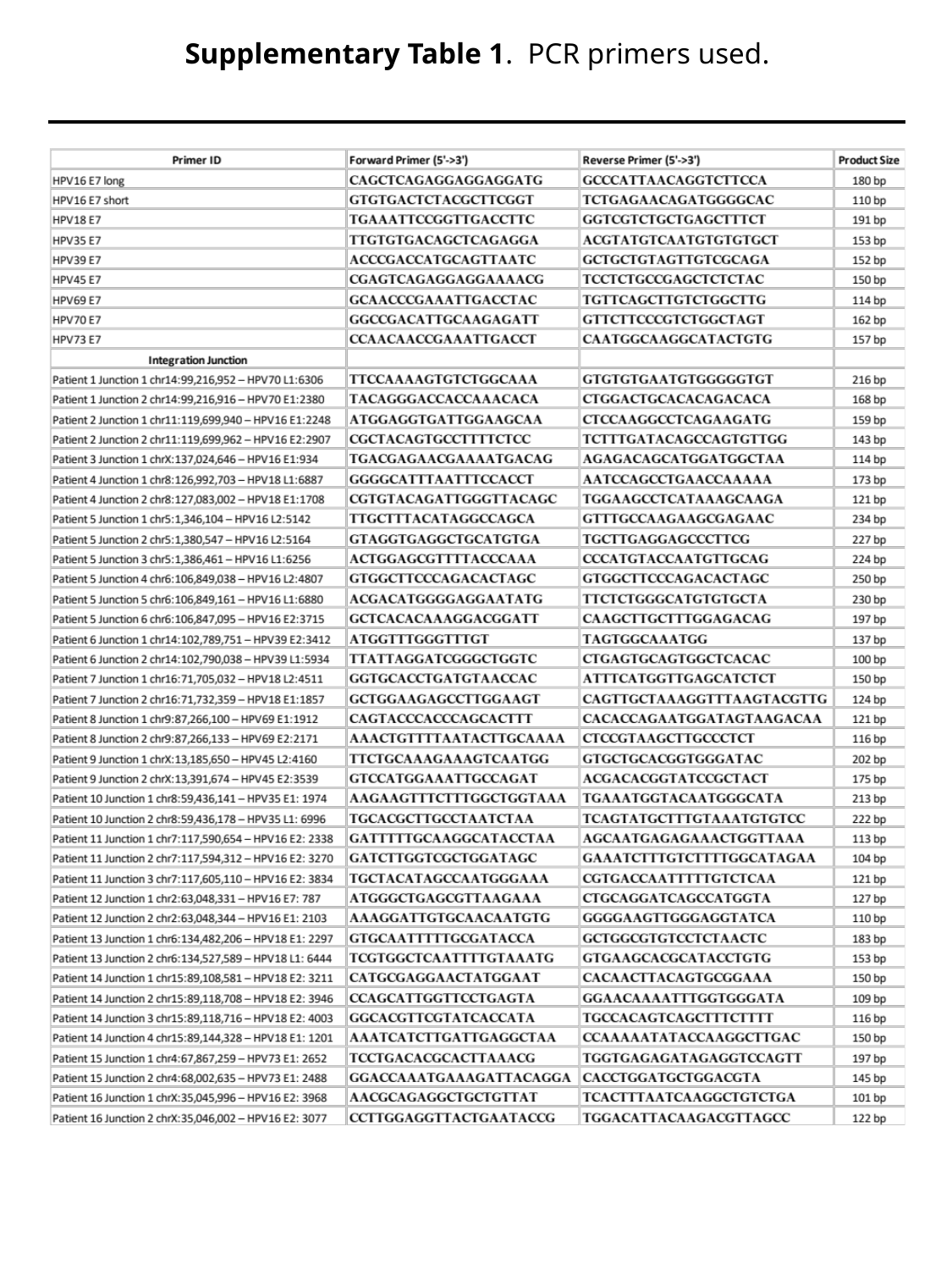

Supplementary Table 1. PCR primers used.

### Slide 2
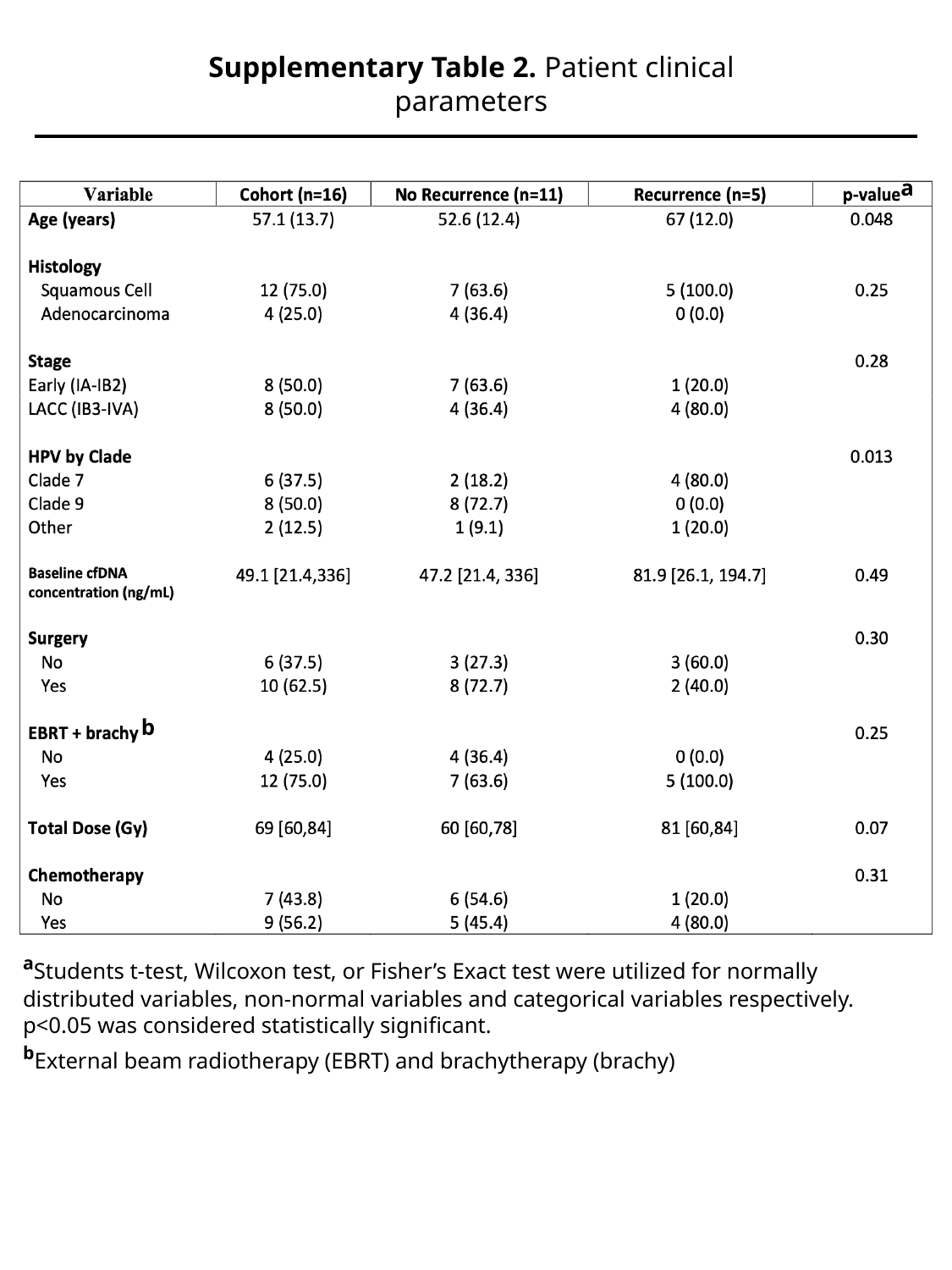

Supplementary Table 2. Patient clinical parameters
a
b
^a^Students t-test, Wilcoxon test, or Fisher’s Exact test were utilized for normally distributed variables, non-normal variables and categorical variables respectively. p<0.05 was considered statistically significant.
^b^External beam radiotherapy (EBRT) and brachytherapy (brachy)
